# Pathophysiological Mechanisms of the Onset, Development, and Disappearance Phases of Skin Eruptions in Chronic Spontaneous Urticaria

**DOI:** 10.1101/2023.11.11.23298424

**Authors:** Sungrim Seirin-Lee, Shunsuke Takahagi, Michihiro Hide

## Abstract

Chronic spontaneous urticaria (CSU) is a typical example of an intractable skin disease with no clear cause and significantly affects daily life of patients. Because CSU is a human-specific disease and lacks proper animal model, there are many questions regarding its pathophysiological dynamics. On the other hand, most clinical symptoms of urticaria are notable as dynamic appearance of skin eruptions (wheals). In this study, we explored dynamics of wheal by dividing it into three phases using a mathematical model: onset, development, and disappearance. Our results suggest that CSU onset is critically associated with endovascular dynamics triggered by basophils positive feedback. In contrast, the development phase is regulated by mast cell dynamics via vascular gap formation. We also suggest a disappearance mechanism of skin eruptions in CSU through an extension of the mathematical model using qualitative and quantitative comparisons of wheal expansion data of real patients with urticaria. Our results suggest that the wheal dynamics of the three phases and CSU development are hierarchically related to endovascular and extravascular pathophysiological networks.

## 1 Introduction

The skin is the most important organ in the human body for maintaining its homeostasis. It has physical and immunological barrier functions to protect the internal organs from the external environment. The skin is involved in the immune system and may develop life-threatening neoplasm, such as cancers and infectious diseases as well as various allergic and/or inflammatory diseases, such as atopic dermatitis, chronic urticaria and psoriasis that severely affect daily life of patients (Leiter et al. 2020; Seidel et al. 2018; Thandi and Whittam 2021; Weidinger et al. 2018; Kolkhir et al. 2022). Although most skin diseases are readily diagnosed by visual observation of skin eruptions, many of them lack appropriate animal models (Kolkhir et al. 2022). Even if animal models are available, many of them cannot precisely replicate correspodning human diseases, making it still difficult to elucidate the whole picture of underlying mechanisms *in vivo* and to develop effective treatments. Thus, it is challenging to fully understand the pathophysiology of skin diseases and find a way to cure by limited clinical data.

Urticaria is one of the most common human skin disorders, affecting at least one in five people during their lifetime (Lee et al. 2017; Maxim et al. 2018). Chronic spontaneous urticaria (CSU), a major subtype of urticaria, is a typical example of an intractable skin disease with unknown cause (Greaves 2014). It is characterized by the appearance of notable skin eruptions called wheals, and is typically accompanied by distressing itching. Wheals of notable shapes recur on a daily basis in patients with CSU, and this significantly affects the quality of life (QoL) of patients’ daily lives over a long timescale, from months to decades (Itakura et al. 2018; Zuberbier et al. 2022). Wheal formation in urticaria is predominantly mediated by histamine which is released from skin mast cells and its action on the microvascular endothelium (Kolkhir et al. 2022; Seirin-Lee et al. 2023). In a certain population of patients with CSU, mast cells can be activated by cross-linkage of IgE against autoallergens, such as interleukin (IL)-24, double stand DNA and thyroid peroxidase, and/or IgG autoantibodies against IgE or the high affinity IgE receptor (Fc*ε*RI) (Maurer et al. 2021). A positive feedback reaction to histamine release from mast cells is suggested to play a role in the expansion of wheals (Bazilai et al. 2017). Antihistamines, the mainstay of treatments for CSU, may be effective in up to 70-80% of CSU patients (Maurer et al. 2011), but their efficacy varies among patients. This indicated that the pathomechanism of urticaria cannot be explained solely by histamine dynamics.

The pathomechanism of wheal formation in CSU has been recently suggested to be involved by basophils in the peripheral blood circulation, blood coagulation, and complement systems (Yanase et al. 2021b; Huang et al. 2020; Rijavec et al. 2021). In our previous studies (Seirin-Lee et al. 2020, 2023), we successfully developed a mathematical model that integrates pathophysiological networks *in vivo* and recapitulated the geometric features of CSU skin eruptions *in silico*. Based on *in silico* experiments with changing parameters of this mathematical model, we obtained five types of eruption patterns with the development of the clinical criteria of CSU eruption geometry (EGe Criteria) and extracted critical pathophysiological networks for each type. Of note, eruptions appeared in almost of 105 patients with CSU were successfully classified into one of these five types based on EGe Criteria by six independent dermatologists. These observations suggest that morphological characteristics of wheals on the skin are predominantly, if not directly, and their related to dynamics of biological molecules and/or cells *in vivo*. Thus, the elucidation of these relationship by mathematical approach is a critical step to proceed forward to understand the overall mechanism of CSU and develop wheal shape-specific treatments.

A daily or mostly daily dynamic phenomenon of the spontaneous appearance and disappearance is a notable characteristic of wheal formation in CSU (Zuberbier et al. 2022). The eruption shapes change, ranging in size from a few millimeters to several centimeters, and in time from several minutes to hours. Clinically, wheal dynamics can be divided into three phases; onset, development, and disappearance. The onset phase is critical as it is most likely linked to the triggering mechanism of the disease. Various patterns of eruptions are appeared during the developmental phase, suggesting the involvement of pathophysiological network in various ways during this phase (Seirin-Lee et al. 2023). However, the detailed mechanism of developing diversity in the patterns of eruptions in CSU remains mostly unexplored. The disappearance phase has received far more less attentions. However, the regulation of this phase could also open a way for therapies to prevent the progress and/or even to prevent the development of wheals. The mathematical model of previous study, Seirin-Lee et al. (2023), was developed based on pathophysiological network assumed to be involved in the onset and development phases, and the disappearance phase of wheal formation in CSU has not been well-explained yet.

In the present study, we focus on more detailed dynamics of wheal formation in CSU and explore the involvement of pathophysiological dynamics in each phase of wheal formation. For this purpose, we choose the mathematical model developed by Seirin-Lee et al. (2023) and analyze it with reconstructing two independent models in the endovascular and extravascular sites. We also suggest a possible mechanism of eruption disappearance in CSU by extending a mathematical model with qualitative and quantitative comparisons of eruption expansion data of urticaria patients. Our results suggest that the dynamics of the three phases of wheal formation in CSU development are hierarchically related to endovascular and extravascular pathophysiological networks and that a global inhibitory mechanism of mast cells is essential in wheal dynamics of the disappearance phase. This study provides a better understanding of the mechanism of CSU by morphological dynamics in wheal formation and its relationship to cellular and molecular dynamics *in vivo*, thereby contributing to accurate diagnostic for individualized treatments decision making in clinical settings.

## 2 Method

### 2.1 Basal CSU model

The pathophysiological structure that develops CSU *in vivo* is assumed as shown in Fig. 1A (Hide and Kaplan 2022; Seirin-Lee et al. 2023). Based on unexplained small stimuli, basophils that randomly adhere to the blood vessel endothelial tissue within capillaries begin to release histamine. The histamine released from basophils enhances the expression of tissue factor on the endothelial cell surface (*N*_1_) which activates the extrinsic coagulation pathway and histamine release from basophils (*N*_2_) (Karasuyama et al. 2009; Stone et al. 2010; Yanase et al. 2021a). This comprises a positive-feedback loop at the endovascular site (*N*_1_ − *N*_2_). At the same time, mast cells stimulated by another resource also start releasing histamine which enhances the expression of tissue factor (*N*_1_) and causes coagulation factors to leak from the blood vessels by creating a gap formation (*N*_3_) (Kurashima and Kiyono 2014; Yanase et al. 2018a,b). This further activates histamine release from the mast cells (*N*_4_) (Yanase et al. 2021a) and induces another positive-feedback loop at the extravascular site (*N*_1_ − *N*_3_ − *N*_4_). Networks *I*_1_, *I*_2_, and *I*_3_ are the networks inhibited by adenosine consequentially produced from adenosine triphosphate (ATP) which is released from basophils and mast cells simultaneously with histamine, and inhibits histamine release and tissue factor expression on endothelial cells (Rudich et al. 2012; Matsuo et al. 2018).

**Figure 1:**
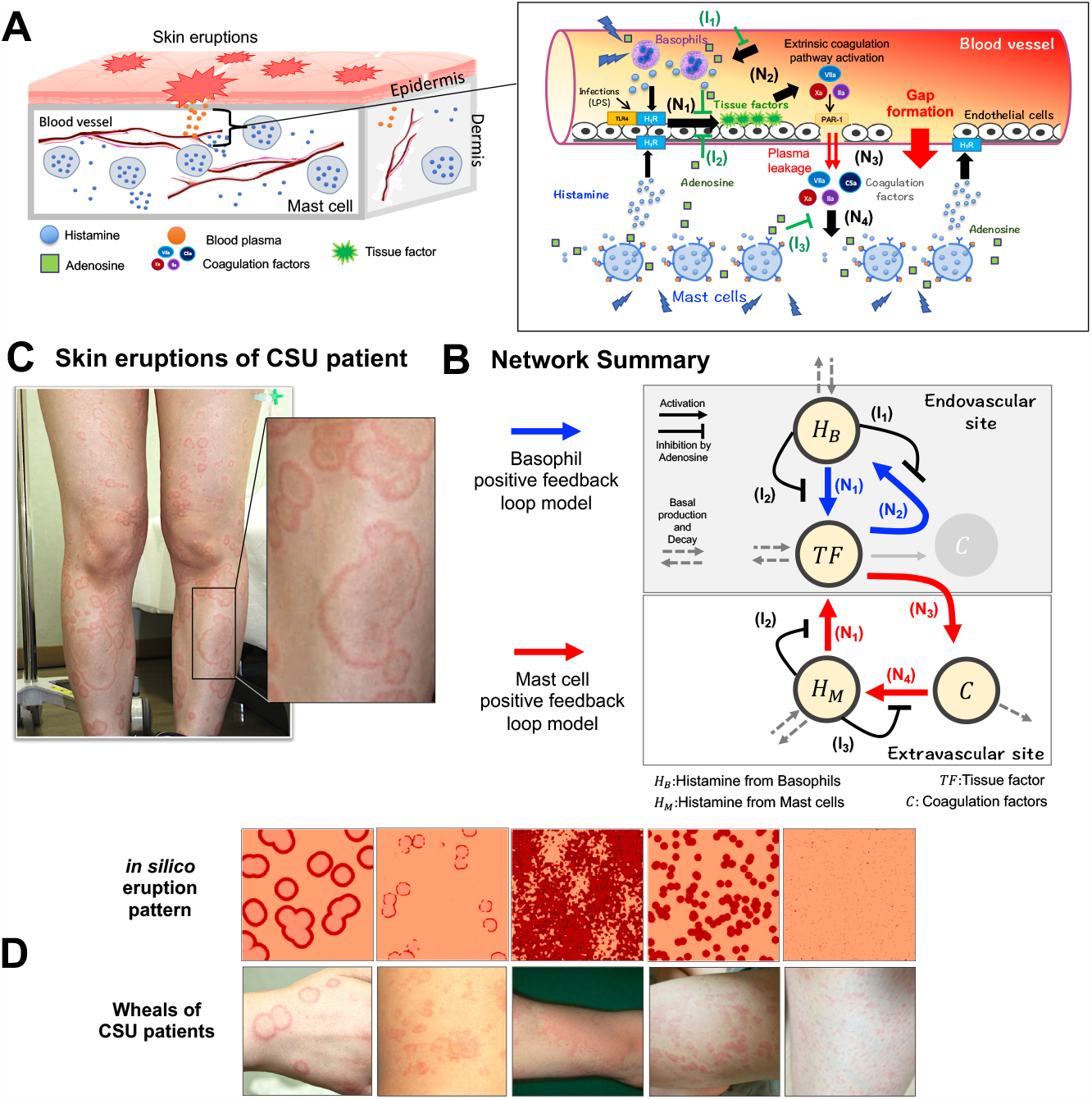
Summary of pathophysiological dynamics of CSU and network. (A) Schematic summary of pathophysiological CSU dynamics and details of endovascular and extravascular sites. (B) Network summary of pathophysiological dynamics in CSU. (A-B) The network *N*_1_−*N*_2_ consists of positive feedback in the endovascular site, and the network *N*_1_−*N*_3_−*N*_4_ consists of positive feedback in the extravascular site. *I*_1_, *I*_2_ and *I*_3_ are inhibition networks by adenosine. (C) Example of skin eruption patterns in a patient with CSU. (D) Regeneration of eruption patterns of CSU using the basal model (1)-(4). The picture was adopted from Seirin-Lee et al. (2022) under a CC-BY-NC-ND 4.0 International license.

The dynamics of CSU development were mathematically modelled in Seirin-Lee et al. (2023) based on *in vitro* experimental data. We recalled and reconstructed these to explore the precise effects of each phase on CSU development. Specifically, the basal model is expressed as:

**[Endovascular site]**

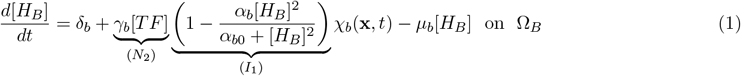

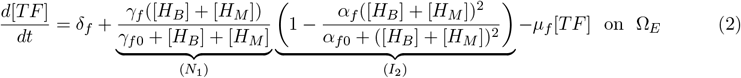

**[Extravascular site]**

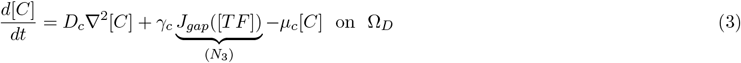

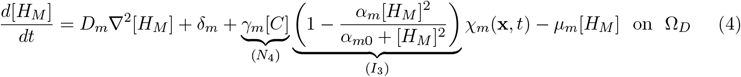

where [*H*_*B*_](**x**, *t*), [*TF*](**x**, *t*), [*C*](**x**, *t*), and [*H*_*M*_](**x**, *t*) are concentrations of histamine released from basophils, tissue factor expressed on vascular endothelial cells, activated coagulation factors leaked from blood vessel, and histamine released from mast cells, respectively. *J*_*gap*_(**x**, *t*) is a function of gap formation estimated from the experiments (Seirin-Lee et al. 2023), and its detailed form is:

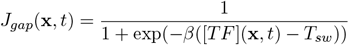

Each *N*_*i*_(*i* = 1, · · ·, 4) and *I*_*j*_(*j* = 1, · · ·, 3) indicate the main physiological networks shown in Fig. 1A and B. Ω_*D*_ ⊂ ℝ^2^ is the region of the dermis, Ω_*E*_ is the vascular endothelial tissue, and Ω_*B*_(⊂ Ω_*E*_) is a random region of the vascular endothelial tissue in which basophils affect the endothelial cells. A certain number of blood vessels are sufficiently and uniformly distributed in the skin and the depth scale at which the vascular reactions involved in wheal formation is negligibly small compared to the scale of the wheal patterns in patients with CSU. Thus, the model is assumed on Ω_*D*_ = Ω_*E*_ = [0, *L*] × [0, *L*] ⊂ ℝ^2^. All parameter values in the model are assumed to be non-negative constants and *T*_*sw*_ *>* [*TF*](0). *χ*_*b*_(**x**, *t*) and *χ*_*m*_(**x**, *t*) are defined as:

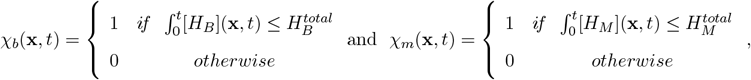

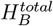 and 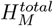 are the total amounts of histamine contained in basophil and mast cells, respectively (Hattori and Seifert 2017).

Because skin eruptions reflect a condition in which substrates leaking from the blood vessels are visible on the skin surface, we define the eruption state function, *S*_*w*_, on the skin (Ω_*P*_ ⊂ℝ^2^) as defined in Seirin-Lee et al. (2023), such that:

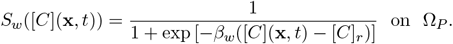

This function directly reflects the qualitative dynamics of the eruption patterns, which is consistent with the dynamics of the main factors in the model.

A previous study by Seirin-Lee et al. (2023) found that the eruption pattern of a CSU can be classified into five types and successfully regenerated *in silico* patterns using the models (1)-(4). Here, we recall the actual eruptions of CSU patients and the representative *in silico* patterns generated by the models of (1)-(4) in Fig. 1C and D.

### 2.2 Basophil/Mast cell positive-feedback loop model

By neglecting the extravascular or endovascular site network loops of either *N*_1_ − *N*_3_ − *N*_4_ or *N*_1_ − *N*_2_, we reconstructed the basal model (1)-(4) as the basophil/mast cell positive-feedback loop model (Fig. 1B). Specifically, we consider the CSU model (1)-(4) with either [*H*_*M*_] ≡ 0 or [*H*_*B*_] ≡ 0. Using these models, we explored which network loops play a key role in each CSU development phase. The former case corresponds to:

**[Basophil positive-feedback loop model]**

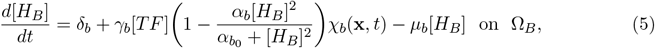

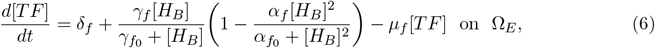

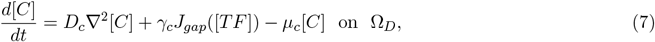

This represents the positive-feedback loop of basophil via vascular endothelium and coagulation factors. The latter case corresponds to the model of

**[Mast cell positive-feedback loop model]**

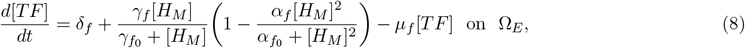

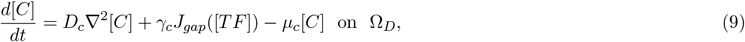

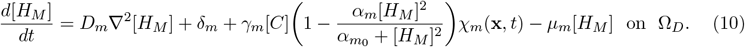

This represents the positive-feedback loop of mast cells via the tissue factor of the vascular endothelium and coagulation factors that leak from the blood vessels.

### 2.3 Histamine-only model of mast cells

To investigate the role of mast cell in developing the CSU more precisely, we reduced the mast cell positive-feedback loop model to a core structure under certain assumptions.

Let us assume that the concentration of tissue factors quickly approaches a steady state, namely, *d*[*TF*]*/dt* ≈ 0, and assume that the diffusion coefficient of coagulation factors is sufficiently slower than that of histamine released from mast cells and that it quickly approaches a steady state, namely, *d*[*C*]*/dt* ≈ 0. Then, models (8)-(9) can be approximated as a system with a sufficiently small *ε* as follows:

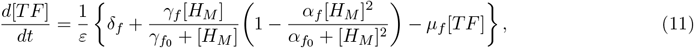

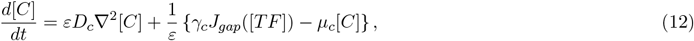

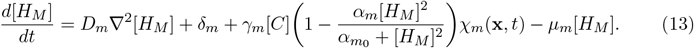

Then, with *ε* → 0, we obtain the following equations from equations (11)-(12):

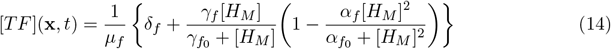

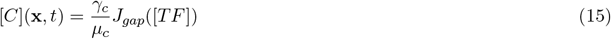

Now, by instituting (11) and (12) into the equation (13), we obtain the reduction system for histamine alone as follows::

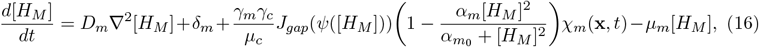

where

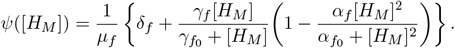

### 2.4 Numerical conditions and parameter values

A numerical simulation was performed by using the alternating-direction implicit (ADI) method (Morton and Mayers 1994) with the zero-flux boundary conditions for [0, *L*] × [0, *L*]. For the initial conditions, we selected a spatially homogeneous state with small perturbations. The detailed forms are as follows:

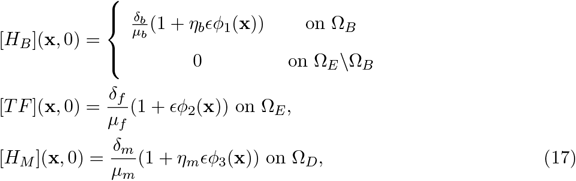

where *ϵ* ≪ 1 and we typically choose 0.01. *η*_*b*_ and *η*_*m*_ are stimulus intensities, and *ϕ*_1_(**x**) is a random variable function of uniform distribution [0, 1] on [0, *L*] × [0, *L*]. Because extravascular coagulation factors cannot leak from blood vessels without gap formation, we assumed that:

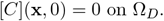

Then, we set Ω_*D*_ = Ω_*E*_ = Ω_*P*_ = [0, *L*] × [0, *L*]. To select a random region of the vascular endothelial tissue in which basophils affect endothelial cells, we selected a specific position for endothelial cells by defining Ω_*B*_ such that Ω_*B*_ = {**x**|*ϕ*_1_(**x**) *> p*} where 0 *< p <* 1. If *p* is close to 1, the position where basophils affect endothelial cells is restricted by a high stimulus point. By changing the value of *p*, the density (denseness or sparseness) of the initial stimuli is represented.

The detailed parameter values were chosen from the original model of Seirin-Lee et al. (2023), in which the representative parameter set was estimated using *in vitro* experimental data. To explore the model dynamics, we tested the parameters in several regions. However, we restricted the inhibition rate to less than 1; namely, *α*_*b*_, *α*_*f*_, *α*_*m*_ ≤ 1, based on the original model development using *in vitro* experiments. The experiments showed that inhibition by adenosine suppressed the histamine release rate but not to a negative value (shown in Figure 2 of Seirin-Lee et al. (2023)).

**Figure 2:**
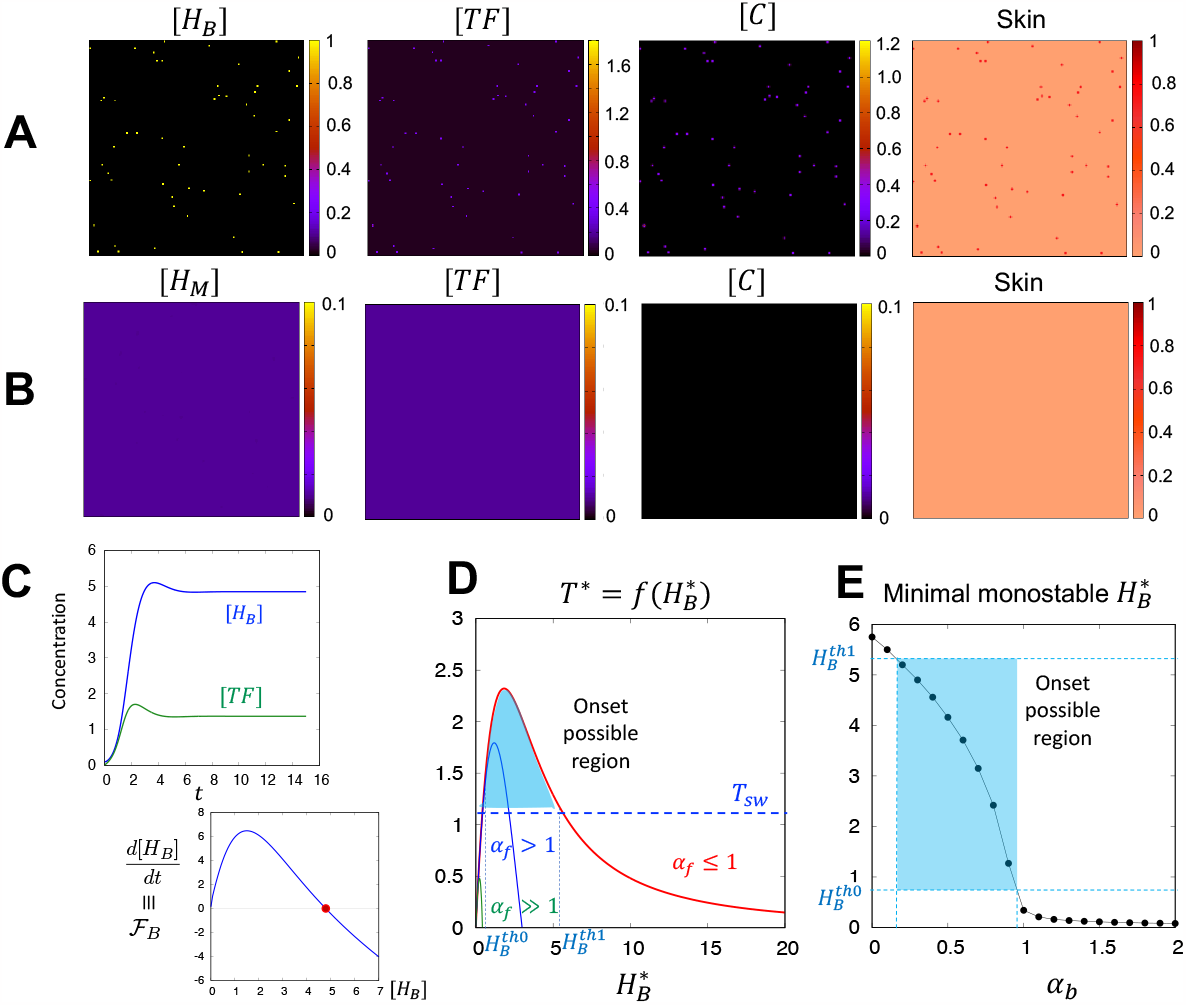
Onset phase of wheal formation in CSU. (A) Simulation results of the basophil-feedback loop model (5)-(7). Dots pattern appears. *μ*_*f*_ = 1, *δ*_*f*_ = 0.01, *γ*_*f*_ = 7, *γ*_*f*0_ = 2.2, *α*_*f*0_ = 8.925, *α*_*f*_ = 1, *α*_*f*_ = 1, *δ*_*b*_ = 0.1, *γ*_*b*_ = 5.2, *α*_*b*0_ = 0.00625, and *α*_*b*_ = 0.335. The values for initial conditions are *η*_*b*_ = 1 and *p* = 0.999. (B) Simulation results of the mast cell positive-feedback loop model (8)-(10). A pattern does not appear. *μ*_*f*_ = 1, *δ*_*f*_ = 0.01, *γ*_*f*_ = 7, *γ*_*f*0_ = 2.2, *α*_*f*0_ = 8.925, *α*_*f*_ = 1, *α*_*f*_ = 1, *δ*_*m*_ = 0.01, *γ*_*m*_ = 5.0, *α*_*m*0_ = 0.0035, and *α*_*b*_ = 0.865. The value for initial condition is *η*_*m*_ = 1000. (C) The simulation result for ODE system (5)-(6) (upper panel) and equilibrium graph of [*H*_*B*_] (lower panel). The same parameter values with (A) were used. The red dot indicates a monostable equilibrium. (D) Possible onset region depending on the inhibition rate *α*_*f*_ of TF expression. *μ*_*f*_ = 1, *δ*_*f*_ = 0.01, *γ*_*f*_ = 7, *γ*_*f*0_ = 2.2, *α*_*f*0_ = 8.925, *α*_*f*_ = 1(red line), 2(blue line), *and*50(green line). The graph is given by the equation (18). (E) Same parameters as those of (D), *α*_*f*_ = 1, *δ*_*b*_ = 0.1, *γ*_*b*_ = 5.2, *andα*_*b*0_ = 0.00625. The value of the monostable equilibrium 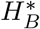was calculated from the equation (19).

### 2.5 Experiment of time courses of annular wheals in urticaria patients

Multiple wheals in the CSU (Fig. 5A) and cholinergic urticaria (Fig. S1) groups with an annular eruption pattern (wheal) were analyzed with the approval of the Ethical Committee for Epidemiology of Hiroshima University, Hiroshima, Japan (approval number: E-1008). The picture data for patient A in Fig. 5A was adopted from Seirin-Lee et al. (2020) under a CC-BY 4.0 International license to confirm the overall changes of wheals with CSU. Wheals that emerged from the patient were recorded using a digital camera over multiple time points. The wheal area was calculated from the number of pixels occupying the target wheal on the digital image, with reference to the number of pixels in the standard length/area.

## 3 Result

### 3.1 Onset Phase: Positive-feedback loop of basophils is crucial for the onset of wheal formation in CSU

We investigated whether CSU onset is due to basophils at the endovascular site or mast cells at the extravascular site. To explore this hypothesis, we considered the following: either the basophil positive-feedback loop model (5)-(7) or the mast cell positive-feedback loop model (8)-(10), with respect to the initial stage (namely, *χ*_*b*_(**x**, *t*) = *χ*_*m*_(**x**, *t*) = 1).

We first tested whether histamine release from basophil is sufficient to induce TF to cause the onset of skin eruptions using the basophil-feedback loop model. We found that the onset of CSU was possible even without the reaction loop of mast cells, although we did not observe any other eruption patterns, except for the dot pattern, in the numerical tests (Fig. 2A).

On the other hand, unexpectedly, we were not able to find any development of eruption patterns in the mast cell-feedback loop model, even for higher levels of the initial stimulus (Fig. 2B). These results indicate that the positive-feedback loop of basophils is crucial for the onset of wheal formation in CSU, and that a stimulus of mast cells may not be a sufficient trigger for the onset.

To confirm our numerical results, we mathematically analysed both models. In the basophil-feedback loop model, the dynamics of CSU onset are determined by the first two ordinary differential equations (ODE) (5)-(6), because the dynamics of coagulation factors are only affected in one way from TF expression. Thus, the possibility of onset can be understood based on the equilibrium conditions of the ODE system at a local point where basophils stimulate endothelial cells. If a positive minimal equilibrium exists which is stable and greater than the initial concentration, onset always occurs. Thus, we first calculate the equilibrium and then investigate the parameter constraints for the existence of a positive equilibrium. By defining the equilibrium concentrations of [*H*_*B*_] and [*TF*] as 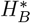 and 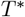, respectively, we obtain:

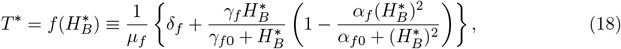

and

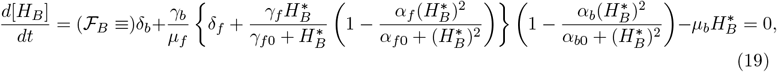

from equations (6)≡ 0 and (5)≡ 0, respectively.

From the equation (19), we see that a unique positive equilibrium exists if *α*_*b*_ *<* 1 and *α*_*f*_ *<* 1 as shown in Fig. 2C (See Appendix A for a more detailed analysis). From a mathematical rather than biological perspective, it follows that if *α*_*f*_ ≫ 1, the equilibrium concentration of [*TF*] becomes lower than the threshold value, below which gap formation cannot occur, namely, *T*^*\**^ *< T*_*sw*_ (Fig.2D (Green line)). That is, CSU did not develop in this case. On the one hand, the onset should only be possible if the equilibrium is within an appropriate range (*T*^*\**^ *> T*_*sw*_ and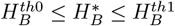). Thus, we next investigate the minimal positive equilibrium 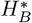 value from equation (19) by changing *α*_*b*_ under the condition *α*_*f*_ ≤ 1. We find a stable minimal equilibrium which satisfies 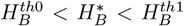 and 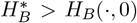 and confirm that onset is possible when *α*_*b*_ ≤ 1 (Fig. 2E (blue shaded region)). Taken together, we conclude that, without parameter sensitivity, onset is possible through the basophil positive-feedback loop.

Finally, we confirm the simulation results shown in Fig. 2B by analysing the mast-cell-feedback loop model in more detail. If the initial TF expression is not sufficient (i.e. [*TF*] *< T*_*sw*_), gap formation does not occur and coagulation factors cannot be leaked from the blood vessels. Thus, the coagulation factors in the extravascular site decay exponentially, which means [*C*] ≈ 0; consequently, the histamine from mast cells will remain in the initial equilibrium state, namely, *δ*_*m*_*/μ*_*m*_ from equation (13). This happened in Fig. 2B.

However, if we assume that [*TF*] *> T*_*sw*_ and the coagulation factors quickly approach the steady state, namely [*C*](**x**, *t*) ≈ *γ*_*c*_*/μ*_*c*_ from equation (9), then, we obtained a single equation for histamine production from mast cells as follows:

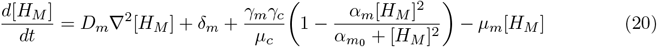

This assumption can be interpreted such that we consider the dynamics of histamine from mast cells under conditions where the expression of tissue factors is sufficiently strong with respect to even a small positive feedback from mast cells. Now, we consider a spatially perturbed stimulus around a constant steady state(≡ *u*^*\**^ *>* 0), namely, [*H*_*M*_](**x**, *t*) = *u*^*\**^(1 + *ϵϕ*(**x**)), where *ϵ* ≪ 1. We can assess that the reaction term in equation (20) has a unique positive equilibrium and the calculation of the linear analysis for the homogeneous steady state confirms a negative eigenvalue (*λ*), such that:

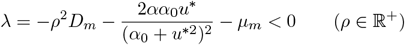

(See Appendix A for more details). This result indicates that all constant steady states are stable without parameter dependence and that the histamine level released from mast cells always reaches equilibrium if TF expression is large enough to cause gap formation. Therefore, the TF expression state is a critical factor for bifurcation to induce onset in the mast cell positive-feedback loop model.

Taken together, locally strong stimulation TF expression via basophil positive-feedback loop induces gap formation and consequently triggers the wheal onset of CSU. However, globally weakly stimulated TF expression via the mast cell-positive-feedback loop may not be sufficient to form a gap, indicating that mast cells may not be fundamental players in the onset of wheal formation in CSU.

### 3.2 Development Phase

In the previous section, we found that gap formation is crucial in the onset phase and the basophil dynamics related to TF expression are critical. However, we could not find any other types of patterns except for dots in the basophil positive-feedback loop model. These observations indicate that endovascular site dynamics play a role in triggering the CSU, while additional mechanisms may be involved during the developmental phase of wheal formation or the developmental phase involves the dynamics of extravascular site. Therefore, we asked the following questions. First, what is the key mechanism that induces the spatial heterogeneity (namely, spatially non-uniform appearance) of skin eruptions during the developmental phase? and, what is the main factor creating the diversity of eruption patterns?

#### 3.2.1 The switch mechanism of gap formation is crucial for generating the spatial heterogeneity of skin eruptions

To investigate the mechanism by which skin eruptions develop spatially and heterogeneously after CSU onset, we focused on the TF-switch role. As we can see in the analysis in the previous section, the level of TF expression is crucial in the onset of CSU, and gap formation plays a crucial role in connecting the endovascular and extravascular sites. In the original CSU model (1)-(4), the gap formation function was estimated by on-off switch type function based on the experimental data of Seirin-Lee et al. (2023). Thus, to understand how the on-off switch type plays a critical role in the overall pathophysiological dynamics of CSU wheal development, we assumed two other types of gap formation functions in addition to the original TF-switch threshold type: a linear and saturation types (Fig. 3A-D, Table 1). A linear type implies that the gap formation occurs in a linearly dependent manner on TF concentration; thus, that there is no threshold-like effect (Fig. 3D, red line). The saturation type represents the effect of a rapid increase and saturation of the gap formation (Fig. 3D, green line).

**Table 1:**
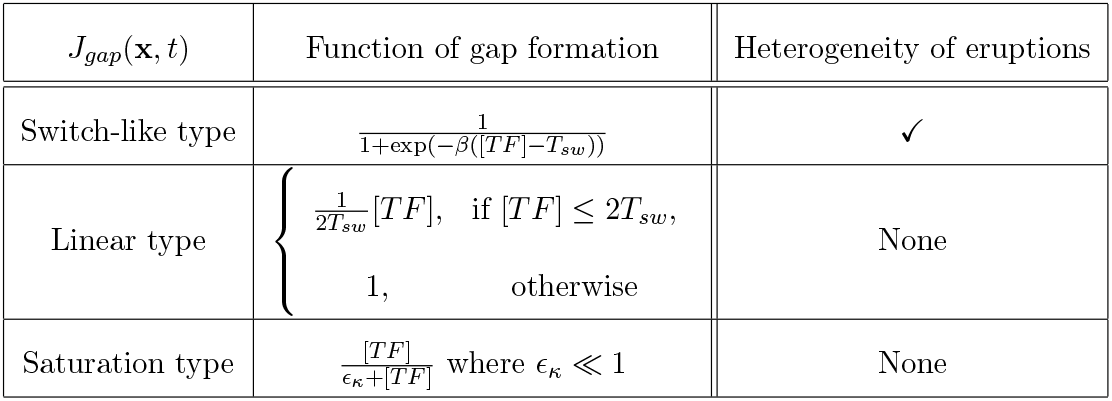
Effect of function types for gap formation on the heterogeneity of eruptions.

**Figure 3:**
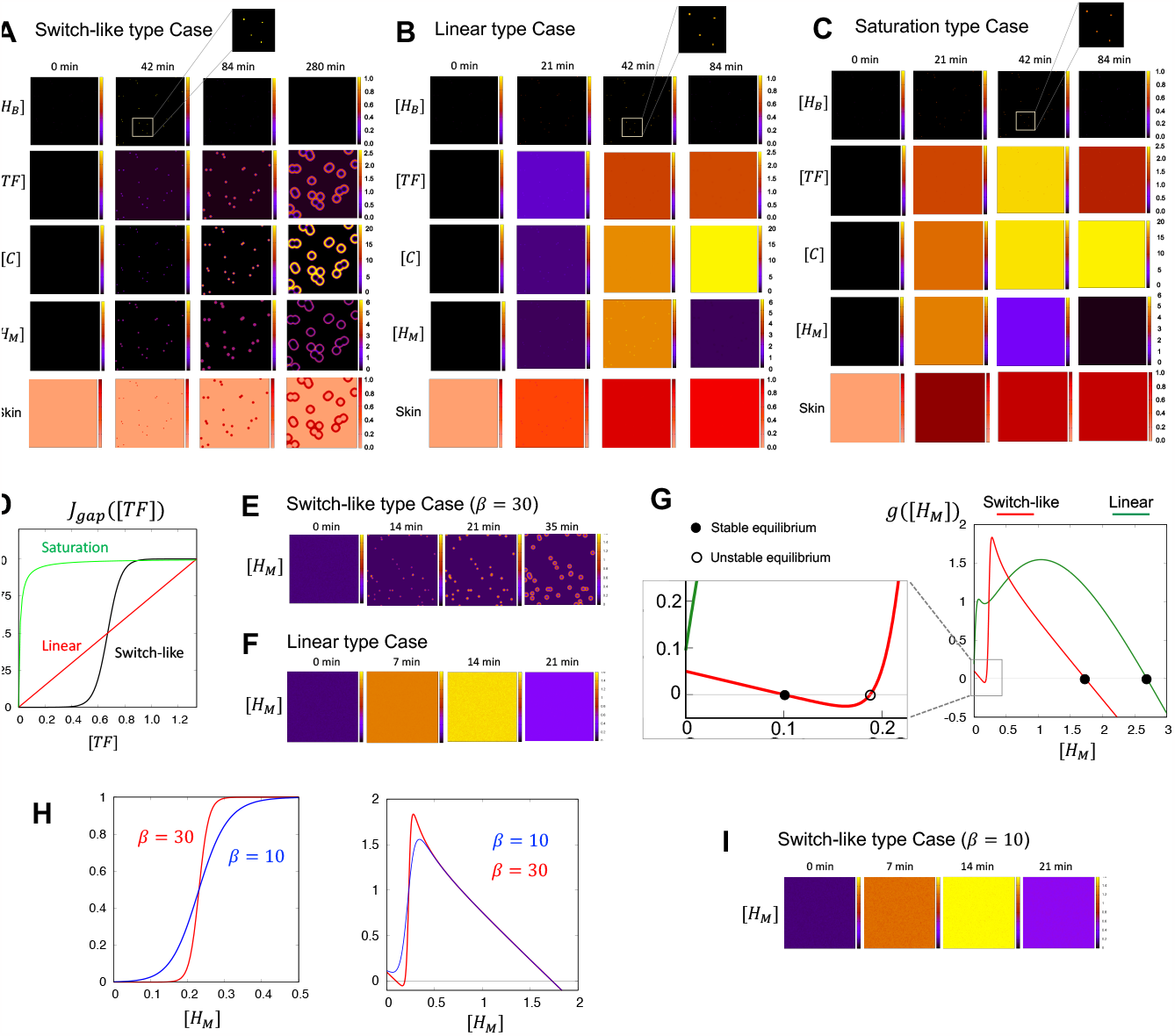
Gap formation functions. (A) Simulation results for the switch-like type in the full model. (B) Simulation results for the linear type in the full model. (C) Simulation results for the saturation type in the full model. (D) The three types of gap formation function. Each *J*_*gap*_([*TF*]) is described in Table 1. *ϵ*_*κ*_ = 0.01. (E) Simulation results for the switch-like type in the reduced histamine-only model. (F) Simulation results for the liner type case in the reduced histamine-only model. (G) Comparison of the reaction functions of the reduced histamine-only model with switch-like and linear cases of gap formation. The switch-like case shows the bistability structure in contrast to the linear case of monostable structure. (H) Gap formation (left panels) and reaction functions (right panels) for the weak (blue line) and strong switch-like cases (red line). (I) Simulation results for weak switch-like cases (blue line in (G)) in the reduced histamine-only model. Representative parameters (Table S1) have been used in (A-C). For (E-I), the parameters are the same as those in Table S1 except for *δ*_*m*_ = 0.1.

We tested the linear and saturation types of gap formation functions (Fig. 3B, C). We found a localized high expression of histamine released from basophil in the endothelial cell tissue, but other factors showed almost spatially homogeneous high concentrations, such as those observed in the case of anaphylaxis. Thus, we were unable to determine the heterogeneity of skin eruptions despite the wheals have been developed. This result indicates that the gap formation dynamics of endothelium are critical for the origin of eruption heterogeneity. The heterogeneous origin of the eruptions was generated by a switch-like system of gap formation.

#### 3.2.2 The development phase is primarily regulated by the positive feedback of mast cell dynamics

We found that the switch-like dynamics of gap formation is critical for the heterogeneous development of wheal. Thus, we hypothesized that the developmental phase may be regulated mainly by the mast cell positive-feedback loop at extravascular site via the vascular endothelium reaction. To confirm this, we reduced the mast cell-feedback loop model (8)-(9) to model (16), which describes only the dynamics of histamine released from mast cells. Let us recall the model (16):

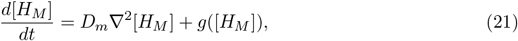

where

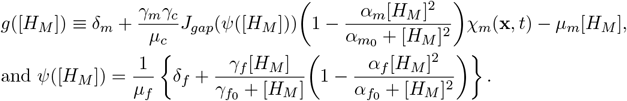

Using the reduction system (21), we first confirmed the state of the pattern development using two gap formation functions:

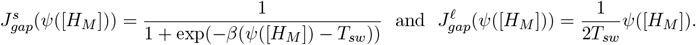

Similar to the full system of (1)-(4), we found a spatial pattern for the switch-like function 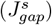 but not a linear gap formation function 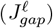, as shown in Fig. 3E and 3F. This result indicates that the spatial heterogeneity of eruption dynamics in the developmental phase can be understood by the dynamics of histamine released from mast cells.

Thus, we explored why switch-like dynamics of gap formation were required for the heterogeneous development of eruptions. We investigated the stability of homogeneous steady states to understand the dynamics of the initial histamine concentration. The linear case exhibited a monostable state (Fig. 3G, green line), indicating that a linear case will present either of the eruptions emerging spatially uniformly because the equilibrium state of histamine 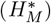 is greater than the initial concentration, namely, 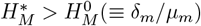(See Appendix B for more detailed mathematical proofs).

However, we found that the switch-like function can create a bistable state (Fig. 3G, red line), which is a critical property that induces heterogeneity in the eruption development. To confirm this, we decreased the switch intensity and found that the bistable property changed to monostable (Fig. 3H). We also confirmed that uniform development (non-patterned state) occurred in the monostable state (Fig. 3I). Therefore, we assume that the switch intensity is sufficiently large, namely, *β* → ∞. Then, we have: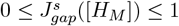,

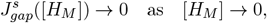

and

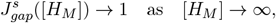

as shown in Fig. 3G (left-hand panel). Then, the following inequality holds:

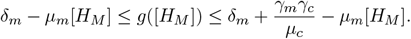

Therefore, the minimal and maximal positive stable steady states (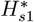 and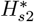, respectively) in equation (28) will be 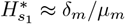 and 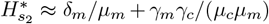based on the comparison theorem (Conway and Smoller 1977). This implies that the system has a bi-stability structure and the initial stimulus size *η*_*m*_*ϵϕ*_3_(**x**) of equation (17) is greater than 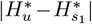 where 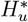 is an unstable equilibrium. [*H*_*M*_](·, *t*) will go to the other stable equilibrium state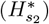. Otherwise, it remained at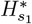. Taken together, a bistable structure with diffusion is likely to cause the development of patterns and the positive feedback dynamics of histamine released from mast cells via switch-like gap formation play a crucial role in wheal development phase of CSU.

#### 3.2.3 Diversity of eruption patterns is largely attributed to the positive-feedback loop of mast cell dynamics

Finally, we investigated the essential mechanisms for creating diverse eruption patterns. As the heterogeneous development of an eruption is regulated by the positive feedback dynamics of mast cells, we hypothesized that the diversity of eruption patterns could be explained by mast cell dynamics. Thus, we numerically tested the types of eruption patterns that appeared in the mast cell histamine-only model (28) by changing the parameter values. Interestingly, we found four types of eruption patterns, which were classified by EGe Criteria developed in Seirin-Lee et al. (2023). The EGe criteria defines five types of eruption patterns: annular, broken-annular, geographic, circular, and dotted. As seen in Fig. 4A, we found four types of patterns except for the dot patterns. We also confirmed that pattern diversity occurs owing to the balance of the three parameters of histamine dynamics from mast cells and TF dynamics in endovascular endothelial cells; the histamine release rate (*γ*_*m*_), histamine decay rate (*μ*_*m*_), and TF activation rate (*γ*_*f*_)) (Fig. 4B). Taken together, the positive-feedback loop of mast cell dynamics is essential in the diversity of eruption patterns, and the balance between activation and inhibition networks in mast cells may be critically involved in eruption diversity.

**Figure 4:**
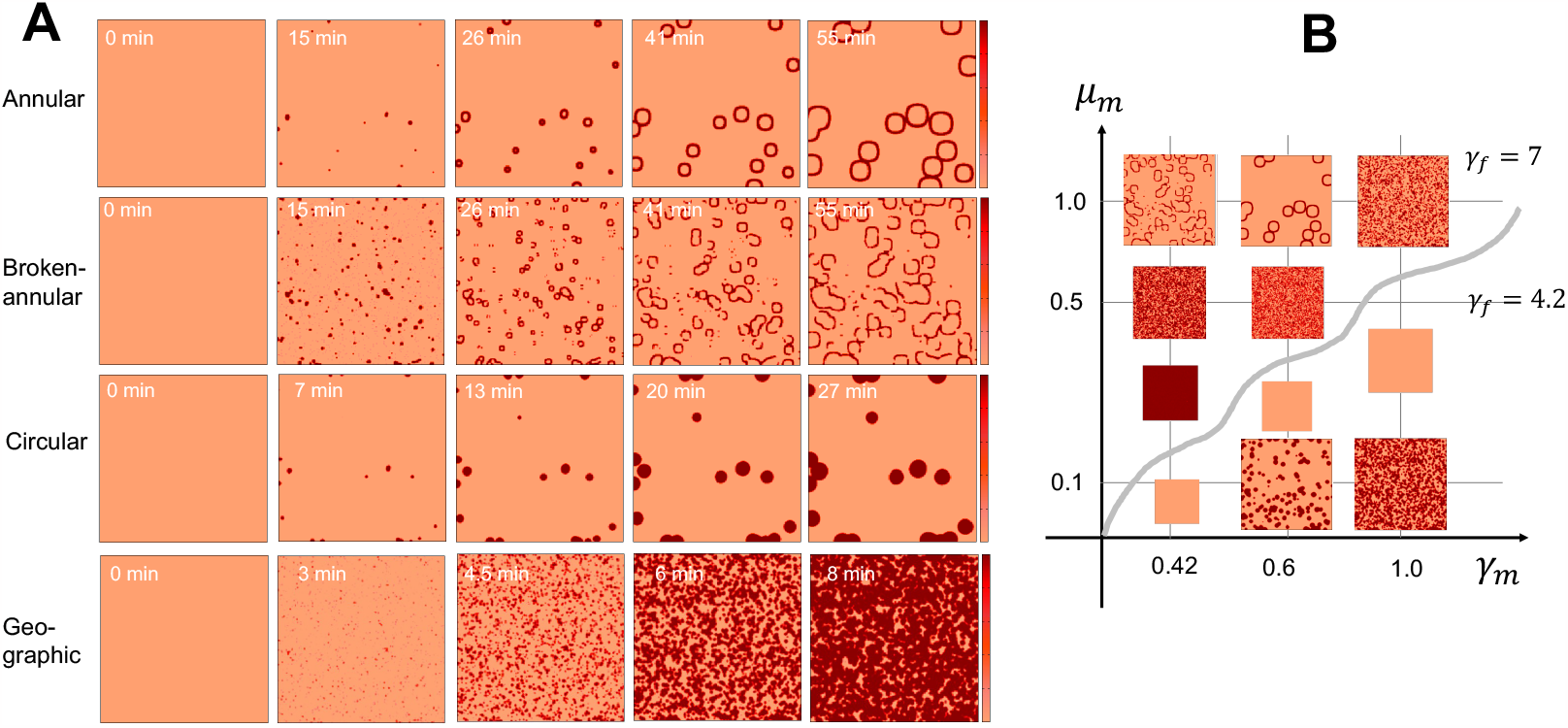
Diverse pattern of eruptions generated by the histamine-only model. (A) Representative simulation examples of eruption patterns. Annular (*γ*_*m*_ = 0.6, *μ*_*m*_ = 1.0, *γ*_*f*_ = 7.0), Broken-annular (*γ*_*m*_ = 0.42, *μ*_*m*_ = 1.0, *γ*_*f*_ = 7.0), Circular (*γ*_*m*_ = 0.6, *μ*_*m*_ = 0.1, *γ*_*f*_ = 4.2), and Geographic patterns (*γ*_*m*_ = 1.0, *μ*_*m*_ = 0.1, *γ*_*f*_ = 4.2) were used and the other parameters are listed in Table S1. (B) Example pattern for each parameter set. The initial condition of this model was given to [*H*_*M*_](**x**, 0) = *δ*_*m*_*/μ*_*m*_ + *η*_*m*_*ϕ*_3_(**x**).

### 3.3 Disappearance Phase: Latent suppression of mast cells may be involved in disappearance of eruptions

One of the characteristics of wheal formation in CSU is the rapid development and disappearance of skin eruptions on a minute or hour scale. Although the pathophysiological mechanism of CSU development was partially or integrally understood in our previous studies, including experiments and mathematical modeling, the underlying mechanism by which urticaria disappears remains elusive. The original models (1)-(4) suggested by Seirin-Lee et al. (2023) successfully regenerated the eruption patterns in the development phase; however, this model failed to regenerate the disappearance phase of the eruptions, except for the dot pattern case. The eruption patterns in this mathematical model generally spread throughout the entire spatial domain if there were pre-simulated mast cells, suggesting that there is a disappearance mechanism separate from the development mechanism of wheals in urticaria. Thus, we chose a mathematical approach to understand the disappearance mechanism. We inferred the mechanism by comparing the quantitative and qualitative dynamics of eruption patterns in a mathematical model with the actual dynamics of wheal patterns in urticaria patients.

We first extracted three features of wheals from the photo data of CSU patients, both quantitatively and qualitatively, as shown in Fig. 5A.

**Figure 5:**
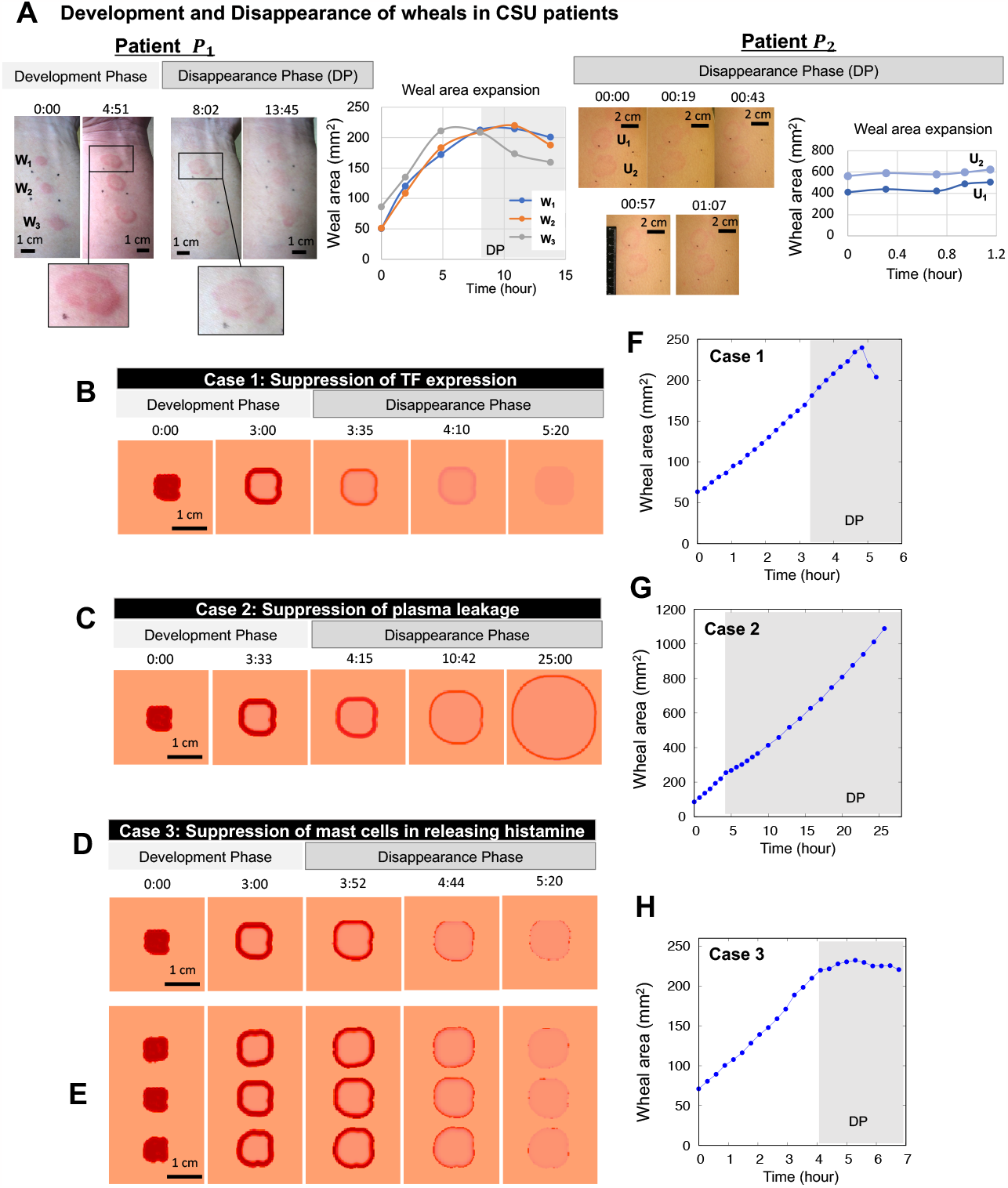
Disappearance phase of wheal formation in CSU. (A) Evolutionary dynamics of wheal (eruption) patterns in a patient *P*_1_ and *P*_2_ with CSU (left panels) and the weal area expansion (right panel). **W**_1_, **W**_2_, **W**_3_ and **U**_1_, **U**_2_ indicate each wheal from the top in each patient figure. The picture of patient A and the data of weal area expansion were adopted from Seirin-Lee et al. (2020) under a CC-BY 4.0 International license. (B-H) *in silico* experiments for the cases of suppression of TF expression, plasma leakage (namely, the intercellular gaps of vascular endothelial cell in blood vessels become small), and mast cells in releasing histamine. (E) Multiple wheals emerge and disappear simultaneously. (F-H) Weal area expansion data calculated from Figs. B, C, D, respectively. The parameter values are listed in Table S1.

i. Wheals that appeared simultaneously will *disappear simultaneously* (the wheals **W**_1_, **W**_2_, and **W**_3_ in left panels in Fig. 5A, Patient *P*_1_).
ii. The boundary of annular wheal becomes *broken* in the disappearance phase (see the enlarged pictures in left panels).
iii. *The wheal expansion almost stops or slightly decrease* in the disappearance phase. (Fig.5A, grey zone (DP) in right graph for patient *P*_1_ and the right graph for patient *P*_2_).

First, based on hints from feature (i), we developed the simplest suppression model that acts uniformly over a wide area after urticaria has progressed to a certain stage:

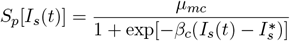

where *μ*_*mc*_ is the suppression rate and *β*_*c*_ is a positive constant that controls the transition state at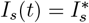. Here, *I*_*s*_(*t*) is defined as:

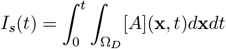

where [*A*] is either a coagulation factor ([*A*] = [*C*]) leaked from the blood vessel or histamine ([*A*] = [*H*_*M*_]) released from the mast cells in the dermis. This modeling formulation implies that a suppression network starts after the development phase of the CSU, and its activation timing is determined not locally for individual wheals, but globally for wheals in the area, depending on the state of each wheal progression. Next, we extend the basal model (1)-(4) by combining the suppression terms as follows:

**Case 1** Suppression of TF expression on vascular endothelial cells: (**S**_**1**_(*t*), **S**_**2**_(*t*), **S**_**3**_(*t*)) = (*S*_*p*_[*I*_*s*_(*t*)], 0, 0).

**Case 2** Suppression of plasma leakage from blood vessels: (**S**_**1**_(*t*), **S**_**2**_(*t*), **S**_**3**_(*t*)) = (0, *S*_*p*_[*I*_*s*_(*t*)], 0).

**Case 3** Suppression of mast cells in releasing histamine: (**S**_**1**_(*t*), **S**_**2**_(*t*), **S**_**3**_(*t*)) = (0, 0, *S*_*p*_[*I*_*s*_(*t*)]).

such that:

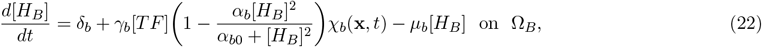

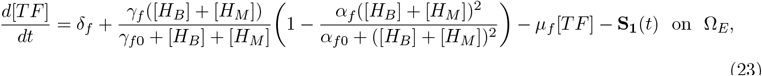

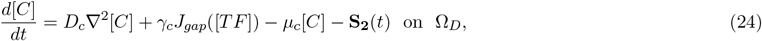

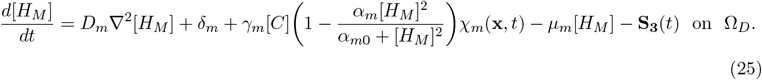

With the disappearance of the phase-combined model (22)-(25), we explored the model case that captured the characteristics of the disappearance phase shown in patients with urticaria in Fig. 5A. Interestingly, we found that only Case 3 succeeded in capturing the wheal evolution characteristics of the CSU patient data, whereas both Cases 1 and 2 failed to explain them (Fig. 5B-H). In Case 1 (Fig. 5B, F), the weal expansion continued in the disappearance phase and a broken pattern was not seen. Features (ii) and (iii) were not found. In Case 2 (Fig. 5C, G), the weal thickness becomes thinner during the disappearance phase, but the wheal patterns did not completely disappear and continuously expended with keeping low concentration of histamine. In contrast, Case 3 satisfied all three features (i)-(iii). The wheal disappeared with a broken annual pattern, and the expansion of wheal almost stops or slightly decreased during the disappearance phase (Fig. 5D, H) (See Fig. S1 in Appendix C for additional clinical data). We also confirmed that Case 3 satisfies wheal patterns disappeared globally (Fig. 5E).

Taken together, we concluded that TF expression or plasma leakage dynamics by gap formation are not be involved, or do not dominantly contribute in the disappearance phase, while the suppression of histamine release from mast cells may play a critical role in the disappearance of wheals in many patients with urticaria.

## 4 Discussion

CSU is an intractable skin disease that involves skin mast cells, basophils, and the blood coagulation and complement systems. IgG autoantibodies to IgE or Fc*ε*RI and/or IgE autoallergy to IL-24, thyroid peroxidase or other endogenous molecules may be involved in the activation of mast cells and basophils (Huang et al. 2020; Maurer et al. 2021; Rijavec et al. 2021; Yanase et al. 2021b). However, due to a lack of proper experimental animal model, the precise mechanism *in vivo* has remained elusive. No individualized medications for each CSU patient has been established, especially to achieve a cure of the disease. A recent mathematical approach (Seirin-Lee et al. 2023) suggested a CSU model, through which the skin eruption dynamics in CSU can be captured. To understand the precise mechanism of CSU development, we focused on the wheal dynamics of CSU by structurally analyzing a mathematical model. We explored the pathophysiological dynamics of wheal formation in three phases: the onset, development, and disappearance phases, and found that the pathophysiological networks play a different role in each phase.

For the onset phase, we found that the basophil positive-feedback loop is critical for evoking the eruptions, but the small stimuli for mast cells were not sufficient to induce wheals in CSU. This observation indicates that the onset of CSU is mainly caused by the reaction of basophils on vascular endothelial cells. The increase of basophil chemoattractant released from skin tissues and/or activation of basophils by autoimmune mechanisms may sufficiently increase the probability of basophils to attach to and activate vascular endothelial cells triggering wheal formation via tissue factor expression. It can also be speculated that the number of basophils in the blood may decrease by moving out of blood vessels to dermal tissues. In fact, the number of basophils in the blood of CSU, especially those who develop with severe symptoms and refractory to treatments, tends to be low (Huang et al. 2020; Rijavec et al. 2021).

During the developmental phase, we found that the mast cell positive-feedback loop was crucial for spatial heterogeneity and various patterns. In particular, we confirmed that the switch-like dynamics of gap formation play a critical role in generating spatial heterogeneity and creating a bistable structure in the mathematical model (Fig. 3). This result suggests that the regulation of vascular permeability by mast cell activation via gap formation dynamics of the vascular endothelium is important. Furthermore, the balance of positive feedback network of mast cells and dynamics of gap formation play an important role in generating various patterns of CSU eruption (Fig. 4B). This also supports the medical understanding that wheal formation in urticaria is primarily driven by mast cells (Zuberbier et al. 2022).

During the development phase, we also found that all four patterns types of eruption reported in the previous study (EGe Criteria, Seirin-Lee et al. (2023)), except for the dot pattern, could be generated in the reduced mast cell model of this study. This indicates that the major, if not all, mechanism of the dot pattern is different from that of the other four patterns. Thus, the efficacy of antihistamine may vary depending on the type of wheals, and that the additional or alternative treatments may be required for patients with dots pattern. The relationship of the efficacy of antihistamine with different patterns of wheals is warranted for future study. In Fig. 4, we often find a high frequency tendency for geographic patterns in the chosen parameter regions. Indeed, the statistical data of patients with CSU on eruption types shown in a previous study, Seirin-Lee et al. (2023), support that geographic patterns were the most dominant patterns found in patients with CSU (Fig. 4B in Seirin-Lee et al. (2023)). The frequency of pattern types should be analyzed more precisely in a future study.

We further confirmed that the positive-feedback loop of mast cell dynamics is essential in the diversity of wheal patterns, and the balance between activation and inhibition networks in mast cells may be critically involved in wheal diversity. This is consistent with the results proposed in the conceptual urticaria model of our previous work, Seirin-Lee et al. (2020). Mathematical insights in this study also showed that bistability based on gap formation function is important in creating spatial heterogeneity in wheal development. A similar structure was found in the previous conceptual model of Seirin-Lee et al. (2020), although the detailed form of the model equation differed. This suggests that an intrinsic and universal mechanism for the diversity of wheals in urticaria is due to mast cell-related dynamics, and that additional networks diverging from the intrinsic structure may give rise to urticarial subtypes.

Finally, our study hypothesized that the disappearance phase of wheals in CSU occurs via a globally independent suppression mechanism followed by the development of wheals and we theoretically validated this by using wheal expansion data of urticaria patients. The *in silico* experiments showed that there may be a mechanism to suppress mast cells from releasing histamine over a wide area where the wheals have developed during the disappearance phase. This suggests that the disappearance mechanism is not a passive dynamic by a local histamine deficiency from stimulated mast cells but an active immune reaction which occurs in a global area over mast cells. Although there is no experimental evidence of the disappearance phase, this hypothesis suggests a novel concept to understand the mechanism of wheal disappearance in that there may exist a global suppression of mast cells after a time lag of wheal development. Mast cells express inhibitory molecules, such as Siglec-8, Siglec-6 and CD200R. The activation of these molecules expressed on mast cells and diminishes histamine release (Metz et al. 2023). The low-affinity IgG receptor (Fc*γ*R2b), which contributes to supra-optimal negative regulation of Fc*ε*RI-induced mast cell activation is another candidate of a player in the disappearance phase (Gast et al. 2018). It would be valuable to prove the precise of pathophysiological network involved in the disappearance phase.

In this study, we explored the overall dynamics of the CSU with respect to the three-phase mechanism of wheal formation by precisely analyzing the CSU mathematical model. Although all the results are based on a mathematical perspective, they suggest a concept of narrowing down the possibility for experimental verification to reveal the precise pathology of CSU. Our theoretical observation of the relationship between the phases of wheal dynamics in CSU and pathophysiological networks may contribute to finding an optimal therapy for patient-specific treatment based on the spatiotemporal dynamics of wheal formation.

## Data Availability

All data produced in the present work are contained in the manuscript

https://github.com/seirin-lee/urticaria-code

## Data availability

All relevant data are included in the manuscript and the supporting information files.

## Code availability

Numerical code is made available online on GitHub (https://github.com/seirin-lee/urticaria-code) and all other data are available from the corresponding author (SSL) on reasonable request.

## Author Contributions

SSL initiated, designed, and executed the research. SSL developed a mathematical model and numerical algorithms for simulations and analyzed data. ST executed the clinical experiment and provided the data. MH provided photographs of patients. SSL wrote the first draft. SSL, ST, and MH revised the manuscript.

## Acknowledgements

This study was supported by the Japan Science and Technology Agency(JST) CREST (JPMJCR2111), and grants to MH from MEXT, Grant-in-Aid for Scientific Research (Grant Number 21K08346).

## Appendix

### A Proofs of stability in onset phase

We consider the two stability problems, independently. One is the equation (19) which has been reduced from the basophil positive feedback loop model (5)-(6). We recall it here;

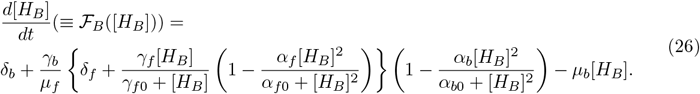

The other is the equation (20)

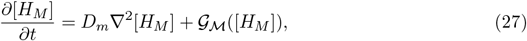

where

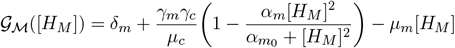

which has been reduced from the mast cell positive feedback loop. Note that the solutions satisfying ℱ_*B*_([*H*_*B*_]) = 0 and 𝒢 ℳ ([*H*_*M*_]) = 0 are constant steady states.

We have the following lemmas.

#### Lemma A.1

*Suppose that α*_*f*_ ≤ 1 *and α*_*b*_ ≤ 1. *Then the minimal positive equilibrium of the equation* (26) *is stable. And the solution started from* [*H*_*B*_](0) *converges to the minimal positive equilibrium*.

<u>Proof.</u> Assume that there exists an equilibrium of *d*[*H*_*B*_]*/dt* = 0 in the equation (26) and let us define the minimal positive equilibrium by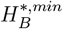. Since ℱ_*B*_(0) *>* 0 and ℱ_*B*_(∞) = −∞,

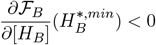

should be hold. Hence, 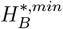 is stable.

Furthermore, from (26), we obtain

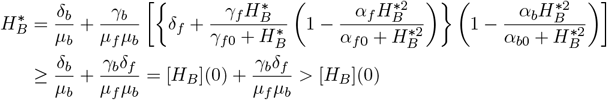

because *α*_*f*_ ≤ 1 and *α*_*b*_ ≤ 1. Since ℱ_*B*_([*H*_*B*_](0)) *>* 0, the solution started from [*H*_*B*_](0) always converges to a stable 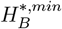.

#### Lemma A.2

*There exists a unique positive constant steady state in the equation* (27) *that is stable for all positive parameters*.

<u>Proof.</u> Since 𝒢 ℳ (0) *>* 0 and

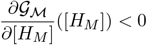

for all positive parameters, 𝒢 ℳ ([*H*_*M*_]) is decreasing in monotone and there exist a unique positive constant steady state. Now, let us denote a constant steady state by [*H*_*M*_](**x**, *t*) = *u*^*\**^. Instituting *z*(**x**, *t*) = *u*^*\**^ + *ϕ*_0_ exp((2*π/L*)**n·x***i* + *λt*), where **n** is a wave mode, into the equation (27) for a standard linear analysis, we have

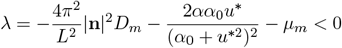

without any constraint of parameter conditions except for the positiveness. Hence, *u*^*\**^ is a stable steady state.

#### Remark A.1

*The reduced model equation* (27) *has been reduced under the condition of sufficient expression of TF. However, if the TF expression is not sufficient, the coagulation factors* [*C*] *will quickly converge to* 0. *Therefore, the reduced model* (27) *becomes*

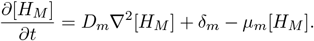

*Thus, u*^*\**^ = *δ*_*m*_*/μ*_*m*_(≡ [*H*_*M*_](**x**, 0)) *is always stable. Thus, onset will not occurs*.

### B Some properties in development phase

We recall the histamine alone model on development phase so that *χ*_*m*_(**x**, *t*) = 1 as the following.

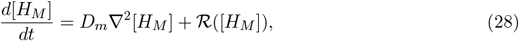

where

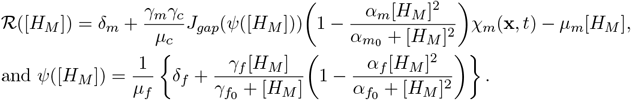

#### Lemma B.1

*Let us define a constant steady state of the equation* (28) *with* 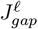 *by* 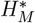. *Then all positive equilibria satisfy* 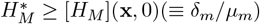 *for α*_*f*_ ≤ 1 *and α*_*m*_ ≤ 1.

<u>Proof.</u> From the reaction terms of (28), we have

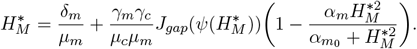

Since we have

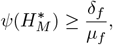

the following inequality is hold;

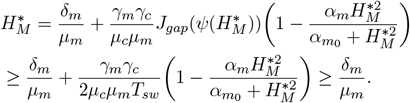

#### Lemma B.2

*Let us suppose that*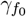 *is sufficiently small such that*

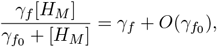

*then the equation* (28) *with* 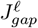 *has a unique constant steady state which is stable*.

<u>Proof.</u> From the equation (28), we can easily check ℛ(0) *>* 0 and ℛ(∞) *<* 0. Thus, the proof is completed if we show that ℛ ([*H*_*M*_]) is a function of monotone decreasing.

The reaction term of the equation (28) denoted by ℛ ([*H*_*M*_]) satisfies

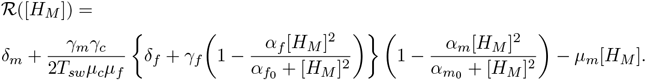

Then, we have

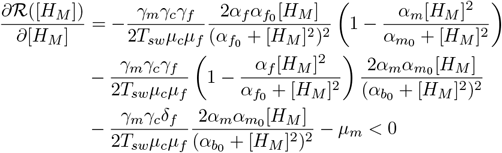

#### Remark B.1

*The two lemmas suggest that if TF expression is constantly induced and the gap formation is linearly dependent on TF expression, there does not exist a spatial eruption patten. Namely, CSU occurs but it is spatially uniform such as the case of Anaphylactic shock*.

### C Additional experiment data and simulation results

The dynamics of wheal expansion was observed in cholinergic urticaria patient and its dynamics shows that the expansion stops in the disappearance phase (Fig. S1 A). We measured the expansion of boundaries with wheal radius for both inner and outer boundaries. The patient data (Fig. S1 A lowest panels) shows that the boundary thickness is narrowing down. On the other hand, *in silico* data (Fig. S1 B) shows that the boundary thickness is narrowing down for the cases of Case 1 and Case 3 in disappearance phase but not in Case 2. Once the disappearance phase enters, the outer boundary stops in contrast that the inner boundary continuously expands as shown in Case 3. Thus, Case 3 show notable shrink of wheal thickness, as seen in both CUS patient in Fig. 5A and cholinergic urticaria patient (Fig. S1 A).

### D Table of parameter values used in simulations

The representative parameter values used in Fig. 3-5 are shown in Table S1.

**Figure S1:**
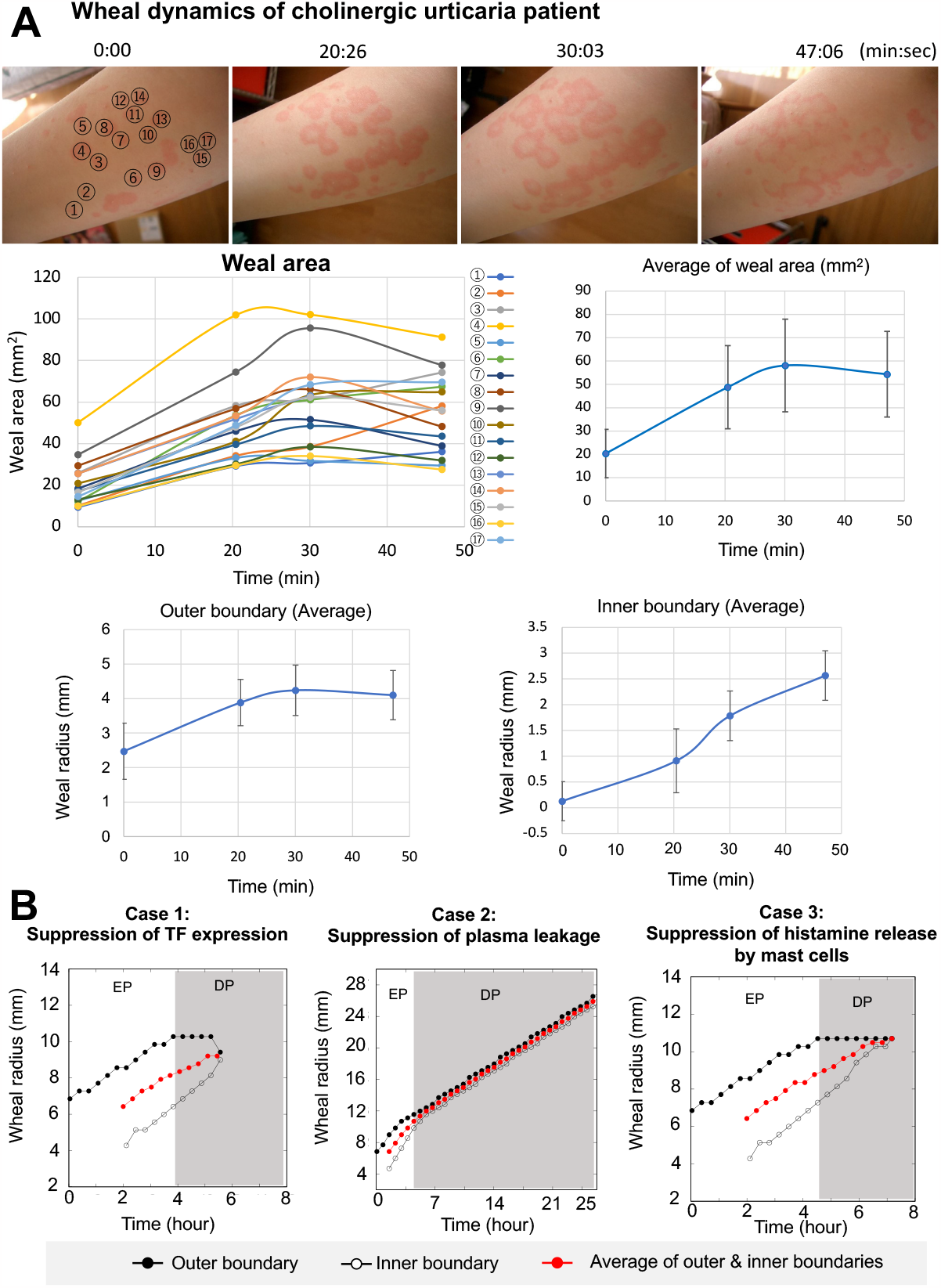
Weal expansion and boundary thickness. (A) Wheal expansion dynamics of patients with cholinergic urticaria. The label of numbers indicates each wheal. The low four panels are quantitative data of wheal area and radius expansion which are calculated from the patient in top photos. The error bar is a standard deviation. (B) *in silico* data for the radius expansion of the wheals calculated in the same simulation of Fig. 5B-D with respect to outer and inner boundaries. Since the wheal expands isotopically, the radius of the wheal was calculated as the distance between the center of the wheal and the point of each boundary when *y*-axis value is the center of the wheal.

**Table S1:** Representative Parameter set used in simulations.

| Parameters | Definition | Value |
| --- | --- | --- |
| $L \times L$ | Spatial length | $1 \times 1$<br>($35.5 \times 35.5 \text{ cm}^2$) |
| $t$ | Time scale | 1.0 (420 sec) |
| $^\dagger D_m$ | Diffusion rates of histamine | $4.7 \times 10^{-6}$<br>($1.412 \times 10^{-5} \text{ cm}^2/\text{sec}$) |
| $D_c$ | Diffusion rates of coagulation factors | $4.7 \times 10^{-6}$ |
| $\delta_b$ | Basal secretion rates of histamine from basophil | 0.1 |
| $\gamma_b$ | Histamine release rate of basophil | 5.2 |
| $^\dagger \alpha_{b0}$ | Parameter affecting the gradient of inhibition rate of histamine from basophil | 0.00625 |
| $^\dagger \alpha_b$ | Maximal inhibition rate of histamine from basophil | 0.335 |
| $\mu_b$ | Basal decay rate of histamine from basophil | 1.0 |
| $H_B^{total}$ | Total amount of histamine of basophil | 600 |
| $\delta_f$ | Basal secretion rates of tissue factor | 0.01 |
| $\gamma_f$ | Maximal increase rate of tissue factor | 7.0 |
| $^\dagger \gamma_{f0}$ | Parameter determining the curve of increasing tissue factor | 2.2 |
| $\mu_f$ | Basal decay rate of tissue factor | 1.0 |
| $^\dagger \alpha_{f0}$ | Parameter affecting the gradient of inhibition rate of tissue factor | 8.925 |
| $^\dagger \alpha_f$ | Maximal inhibition rate of histamine from basophil | 1.0 |
| $\gamma_c$ | Leakage rate of coagulation factors from blood vessel | 20.0 |
| $\beta$ | Parameter determining the stiffness of switching | 20.0 |
| $T_{sw}$ | Switch value of tissue factor | 0.67 |
| $\mu_c$ | Basal decay rate of coagulation factors | 1.0 |
| $\delta_m$ | Basal secretion rates of histamine from mast cell | 0.01 |
| $\gamma_m$ | Histamine release rate of mast cell | 5.0 |
| $^\dagger \alpha_{m0}$ | Parameter affecting the gradient of inhibition rate of histamine from mast cell | 0.00004375 |
| $^\dagger \alpha_m$ | Maximal inhibition rate of histamine from mast cell | 0.865 |
| $\mu_m$ | Basal decay rate of histamine from mast cell | 1.0 |
| $H_M^{total}$ | Total amount of histamine of mast cell | 600 |
| $\mu_{mc}$ | Suppression rate in disappearance phase | 10.0 |
| $\beta_c$ | Parameter determining the transition state of suppression level | 0.05<br>0.05 |
| $I_s^*$ | A threshold concentration of coagulation factor to start a suppression | 350<br>350 |
| $\beta_w$ | Parameter determining the stiffness of wheal state function | 1.0 |

<sup>†</sup> A parameter derived from fitting experimental data in Seirin-Lee et al. (2020) and Seirin-Lee et al. (2023).

